# Language Profile of Posterior Cortical Atrophy: A Comparative Study with Alzheimer’s Disease Variants

**DOI:** 10.1101/2024.12.21.24319481

**Authors:** Linshan Wang, Marie-Anne St-Georges, Monica Lavoie, Raffaella Migliaccio, Maxime Montembeault

## Abstract

**Objective:** This study investigates language impairments in early-stage posterior cortical atrophy (PCA) patients, examining five language subdomains to resolve existing controversies and gaps in the literature.

**Methods:** Participants diagnosed with posterior cortical atrophy (PCA; n=105), typical Alzheimer’s disease (tAD; n=105), logopenic variant primary progressive aphasia (lvPPA; n=116) and healthy controls (HC; n=165) were selected from the National Alzheimer’s Coordinating Center (NACC) database. We utilized language tests from the Uniform Data Set and Frontotemporal Lobar Degeneration Module to assess different aspects of linguistic ability, including verbal fluency, reading, naming, semantics and repetition.

**Result:** Our findings revealed a global decline in visual and non-visual language functions among PCA patients compared to HC, with no spared domains. Furthermore, we investigated specific language errors in reading and sentence repetition, and we found that PCA patients committed a mix of phonological, semantic and word omission errors. They were more impaired on irregular vs. regular word reading and more impaired on verb vs noun naming. Overall PCA patients showed less severe language deficits than lvPPA, except in single word comprehension and verb naming, where the opposite pattern was found. They also showed more impaired visual language impairments and similar non-visual language impairments in comparison to tAD.

**Discussion:** These findings highlight that language impairments in PCA extend beyond visual deficits, playing a key role in its clinical presentation. Recognizing these language issues is essential for differentiating PCA from tAD and lvPPA, where distinct patterns of impairment help refine diagnosis.

## 1. Introduction

Posterior cortical atrophy (PCA) is a neurodegenerative disorder characterized by significant visual difficulties without the presence of primary ophthalmologic causes^1,2,3^. Because this clinical syndrome is most often associated with Alzheimer’s disease (AD) pathology (beta-amyloid plaques and tau tangles), it is sometimes referred as the visual variant of AD^4,5^. As its name suggests, PCA primarily affects the posterior part of the brain including, the parietal, occipital lobes, and posterior temporal lobes^6,7^. The latest diagnosis consensus criteria by Crutch et al. 2017 states that at least three of the following symptoms are present and impact activities of daily living: space perception deficit, simultanagnosia, object perception deficit, constructional dyspraxia, environmental agnosia, oculomotor, dressing and limb apraxia, optic ataxia, alexia, left-right disorientation, anterograde amnesia, acalculia, limb apraxia, prosopagnosia, agraphia, homonymous visual field defect, and finger agnosia. The PCA cognitive profile involves the relative preservation of other cognitive domains such as executive functioning, episodic memory, behavior/personality and nonvisual language functions.

However, non-visual symptoms have been frequently reported in PCA patients, and even if they are not at the forefront of the clinical syndrome, they can be crucial in the differential diagnosis, disease monitoring and intervention in patients with PCA. In fact, working memory^8,9^, executive functions^10^, episodic memory^11^, and social cognition^12^ deficits have been reported in PCA. Additionally, psychiatric symptoms like apathy, depression, anxiety, and irritability are common^13,12^. Zooming in on the domain of speech and language, alexia and agraphia are recognized features and are part of the consensus criteria in PCA. While reading and writing deficits are well documented in the literature and are largely attributed to the visual symptoms central to PCA^14,15,16^, these impairments can also extend to cognitive domains that are independent of visual function, suggesting that they may be more multidimensional than previously anticipated^17^. Nonetheless, very few studies have carefully investigated speech and language in these patients, even if a recent meta-analysis shows that at least 32% of PCA patients have non-visual language and speech deficits^4^.

Confrontation naming is probably the most frequently investigated language domain in PCA, and all studies are consistent in demonstrating a naming impairment in this population^18,19,20^. While object perception deficits can contribute to these impairments, word-finding problems have also been noted in PCA in non-visual tasks such as auditory naming^21^. Interestingly, phonological errors have been observed in naming tasks in more than 55% of PCA patients^22^. Other lexico-semantic impairments have also been reported in PCA, with patients demonstrating difficulties in verbal fluency, word-picture matching and pictorial semantic associations^20,18^. These lexico-semantic deficits in PCA have also been observed in non-visual spontaneous speech and in synonym judgment tasks where words are presented in both spoken and written forms^20,18^ (Although see Rezaii et al. 2024 for conflicting results). Verbal short-term memory deficits, including in sentence repetition, are also documented in PCA^24,20^. Other, less frequently studied language deficits in PCA include grammatical comprehension, auditory input processing—particularly in tasks such as minimal pairs discrimination, prosody pair discrimination, and linguistic prosody (stress discrimination)—as well as speech rate in spontaneous speech^20^.

Given the word-finding, verbal short-term memory (including sentence repetition) and phonological difficulties reported in PCA, many studies have drawn parallels with another atypical AD variant, the logopenic variant of primary progressive aphasia (lvPPA)^25,26,27^. These three deficits are central to the core diagnostic criteria for lvPPA^28^. Furthermore, while PCA causes atrophy in the bilateral temporoparietal-occipital regions, lvPPA affects the left temporoparietal junction^29^. These similarities have led some researchers to coin the term ‘logopenic syndrome’ to describe the language profile of PCA patients, which could be seen in as many as 8 of 9 PCA patients^25^. Crutch et al. (2017) also introduced the PCA-plus label, which can be applied to PCA patients who exhibit additional language symptoms such as anomia and impaired repetition. A recent study involving AD biomarker-confirmed lvPPA and PCA patients found that only 25% of them presented with a purely language or visual AD profile, emphasizing the frequent syndromic overlap in atypical AD and the resulting complexities in achieving accurate categorical diagnoses^30^. Despite similarities between lvPPA and PCA, important distinctions in language deficits have also been highlighted, the main one being that PCA patients generally exhibit milder language impairments. Specifically, PCA patients are less impaired in tasks involving linguistic prosody, word and sentence repetition, lexical and grammatical comprehension, picture and auditory naming, and verbal fluency, in comparison to lvPPA^20,31^. PCA patients also show fewer phonological errors^22^ and better spontaneous speech abilities^20^ compared to lvPPA. Conversely, PCA patients are more impaired in action naming tasks^25^.

Numerous limitations and gaps exist in the literature on speech and language deficits in PCA. Beyond the work of Crutch et al. (2013), no study has provided a comprehensive report across all language domains in PCA. Most studies to date have been case studies or small group studies with 20-25 patients or fewer. Additionally, very few studies have gone beyond total scores to conduct error profile analyses^22^, which have proven valuable in other populations for understanding the cognitive mechanisms underlying clinical impairments^32^. Finally, controversies persist regarding the differences in language profiles between PCA and other AD populations, such as lvPPA and even tAD. Although tAD patients have been reported to exhibit language profiles similar to those of PCA, more comprehensive studies are needed to clarify these relationships^33,34,25,18,21,31,21^.

The current study aims to conduct a thorough assessment of a large sample of early-stage PCA patients across five language subdomains. We hypothesize that PCA patients will demonstrate decreased linguistic performance not only in visual language functions (reading, semantics, and picture naming) but also in non-visual language functions (fluency and verbal short-term memory). Secondly, we intend to explore errors occurring in language tasks (reading and repetition) beyond overall scores, which we believe could aid in the early diagnosis of PCA. We anticipate that PCA patients will make a mix of phonological and semantic errors. Thirdly, we aim to compare PCA with two other major AD conditions, tAD and lvPPA. We hypothesize that PCA will exhibit less severe impairment than lvPPA but more severe than tAD.

## 2. Methods

### 2.1 Participants

The participants in this study were recruited from the National Alzheimer’s Coordinating Center (NACC) database and included cognitively unimpaired older individuals (healthy controls, HC), as well as those diagnosed with PCA, tAD, and lvPPA. Diagnoses were made based on established research criteria^35,36,28^. This study used data from 46 ADRCs for Uniform Data Set (UDS) visits conducted between September 2005 and December 2022. Written informed consent was obtained from all participants or their families.

Participants in the PCA and lvPPA groups were required to have at least one language test result from either the Uniform Data Set (UDS) or Frontotemporal Lobar Degeneration (FTLD) Module language measures. In contrast, individuals in the HC and tAD groups were required to have completed all language tasks. This approach was adopted to balance the sample sizes, since the number of HC and tAD patients considerably outnumbered those of PCA and lvPPA in the dataset. We then excluded all participants who were not English speakers. Finally, patients who scored two or higher on the Global CDR® scale were excluded from the study. With the above criteria, our study included 105 individuals diagnosed with PCA, 105 with tAD, 116 with lvPPA, and 165 HC.

### 2.2. Language tests

All behavioral and social cognition assessment tools from the Uniform data set version 3 and the Frontotemporal Lobar Degeneration Module (FTLD-MOD) were used for this study^37,38,39,40^.

#### 2.2.1. UDS language measures

*The Category Fluency test* measures the ability to generate words belonging to two categories, animals and vegetables, within one minute. Raw score was calculated by counting number of unique responses named within the time limit. Total score for category fluency test was calculated by adding up animal and vegetable categories raw scores.

*The Phonemic Fluency Test* evaluates an individual’s ability to generate words beginning with specific letters, “F” and “L”. Participants are given one minute and asked to produce as many words as they can starting with the designated sound. Raw score was calculated by counting number of unique responses named within the time limit. Total score for category fluency test was calculated by adding up ‘F’ and ‘L’ raw scores.

*The Multilingual Naming Task* involves presenting participants with 32 pictures and asking them to name the presenting pictures^41,42^. Total score is calculated by number of items named correctly within the time limit and a maximum score is 32.

*Number Forward Span Test* measures short-term verbal memory. In this task, participants are presented with a sequence of numbers and are required to repeat them in the same order. The length of the sequence gradually increases from 3 to 8. Total score is calculated by number of correct responses out of 14 items.

#### 2.2.2. FTLD**D**MOD language measures

*Regular Words reading* assess an individual’s ability to read and pronounce regular words. Participants were given a list of 15 words that can be decoded using regular phonetic rules, such as “rope” or “cane”. Total score is calculated by number of correct responses, and a maximum score is 15. Number of phonemic errors (e.g. ‘boss’ for ‘ball’) were also recorded and considered.

*Irregular Words Reading* evaluates an individual’s ability to read and pronounce irregular words that do not follow regular phonetic rules, such as “aisle” or “choir”. Total score is calculated by number of correct responses, and a maximum score is 15. Number of phonemic and regularization errors (e.g. ‘sig’ for ‘sigh) were recorded and considered.

*Sentence Reading* is a task that assesses an individual’s ability to read 5 sentences accurately and fluently. Participants are presented with sentences of varying difficulty levels and asked to read them aloud. Total score is number of completely accurate sentences out of 5 items. Number of omission errors, semantically related errors and phonologically related errors were reported respectively as well.

*The Semantic Word-Picture Matching Test* measures an individual’s ability to match spoken words with the corresponding picture (out of four choices). Total score is calculated by number of correct responses out of 20 items.

*The Semantic Associates Test* is used to assess an individual’s ability to associate items utilizing semantic knowledge. Participants were presented with two pairs of pictures, and they are asked to select the pair that are related (e.g. lion-meat vs lion-corn). The two categories for the pairs are animals and tools and there were 8 items for each. Total score is number of correct responses out of 16 items.

*Sentence Repetition* is a task used to assess an individual’s ability to listen to and repeat sentences. Total score is number of completely accurate sentences out of 5 items. Additionally, number of omission errors, semantically related errors and phonologically related errors were recorded.

*Noun and Verb Naming* evaluates an individual’s ability to produce the names of common objects and action items. Participants are shown 16 drawings of objects, and 16 drawings of action items, they are asked to name the depicted items. Total score for each item type is number of accurate responses out of 16 trials each for noun and verb (Thompson et al., 2012).

### 2.3. Statistical Analysis

Data-preprocessing and statistical analyses were carried out using R version 4.3.1. Demographic features including age, level of education, disease duration, MocA score and total CDR® Dementia Staging Instrument score. These features were compared using simple linear regression model to investigate the presence of demographic and disease severity differences between the groups. Proportion of females and males between each group was compared using Chi-square test. To compare the performance of the groups on each language measures and error types, analyses of covariance (ANCOVA) controlling for age, sex and education were used. Post-hoc comparisons were done using Tukey. Finally, to explore within-test dissociations, we applied two-way ANOVA analyses on noun & verb naming and regular & irregular word reading, between HC and PCA group, controlling for age, sex and education.

## 3. Results

### 3.1. Demographics

Demographic characteristics for the four groups are presented in Table 1. A total of 165 HC, 105 PCA, 105 tAD and 116 lvPPA patients were included. The four groups were matched on age and years of education. However, there were significant differences in sex between HC and lvPPA and PCA and lvPPA (more males in the lvPPA group). All analyses were controlled for age, sex and education. As expected, the three patient groups showed significantly higher disease severity compared to the HC. There were no significant differences between PCA and tAD/lvPPA on MoCA score and disease duration. However, PCA patients had significantly higher CDR® scores than lvPPA patients. Most of PCA patients with available AD biomarkers exhibited positivity for beta-amyloid (Positive/Negative/Unknown: 46.7%/4.7%/48.6%) and tau (35.2%/3.8%/61.0%).

**Table 1.**
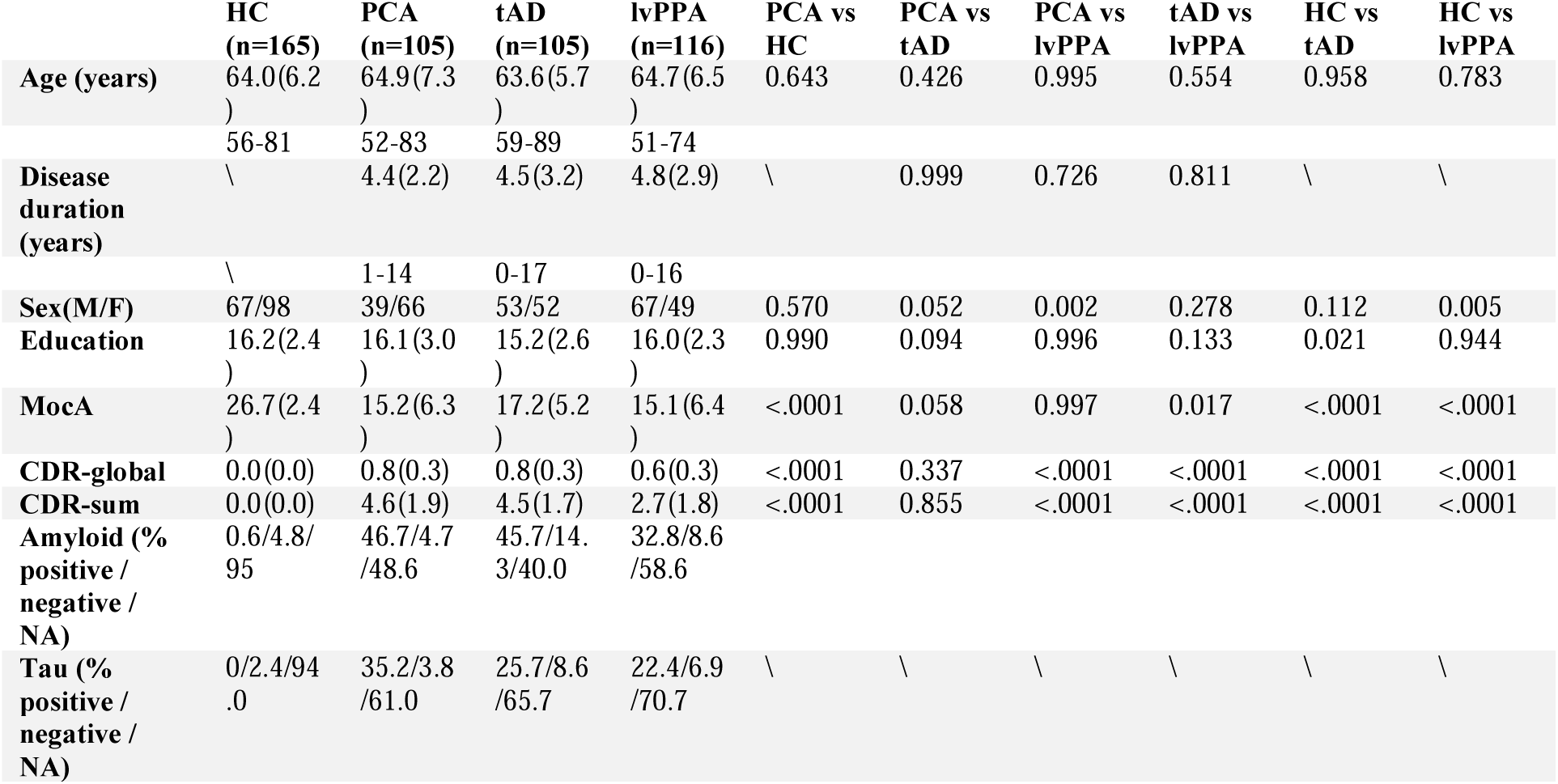
Demographic and clinical description of the participants.

### 3.2. Language measures

Total scores and between-group comparisons for language measures are presented in Table 2.

**Table 2.**
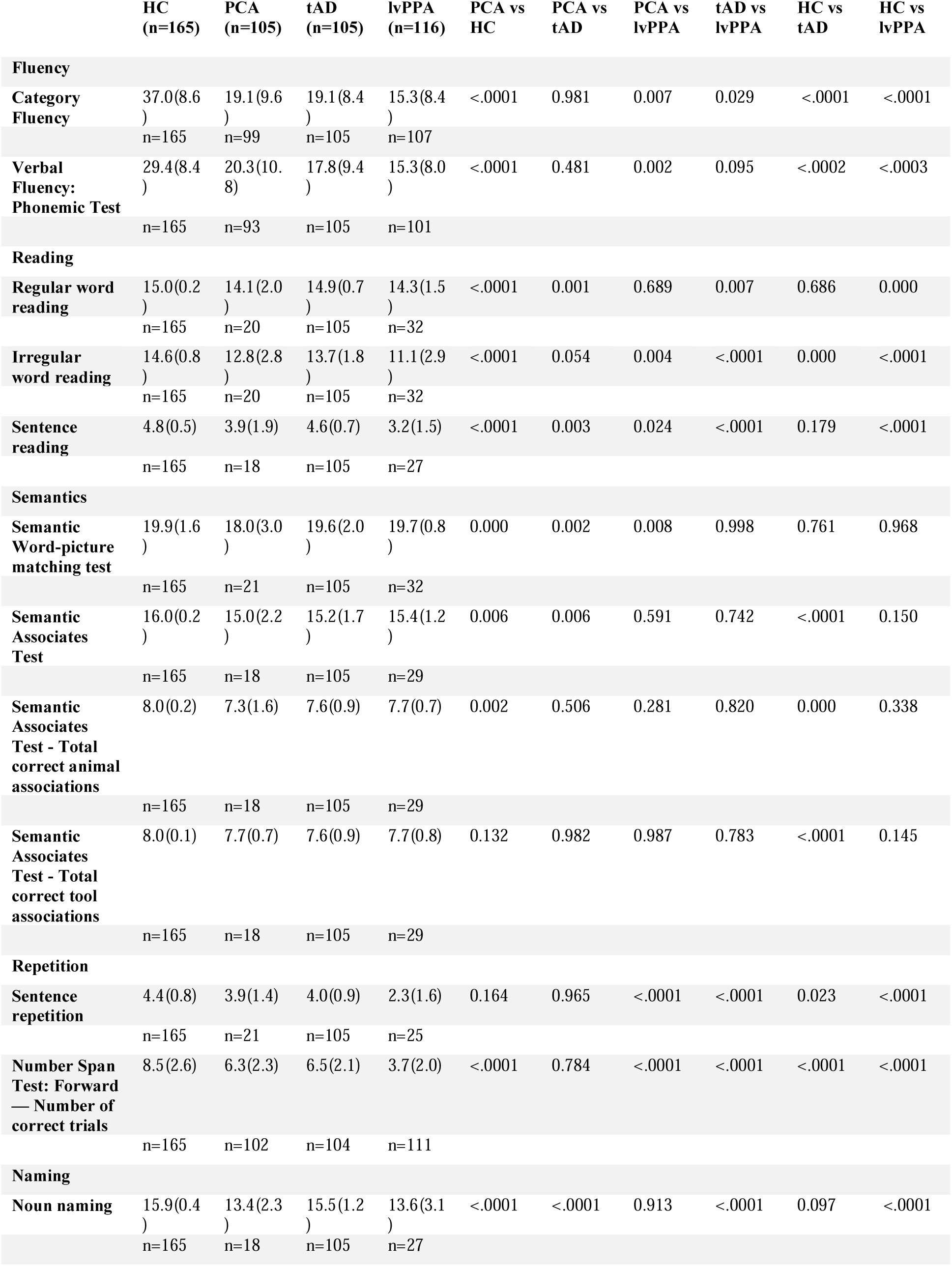
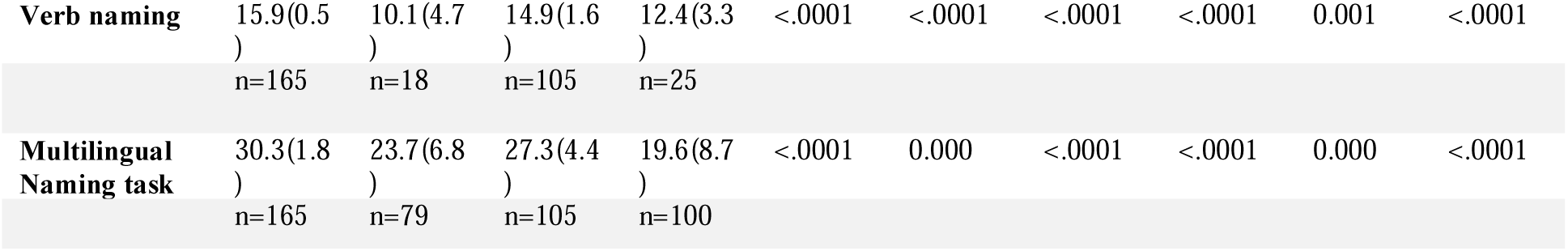
Between-group comparisons on language tests total scores.

|  | HC<br>(n=165) | PCA<br>(n=105) | tAD<br>(n=105) | lvPPA<br>(n=116) | PCA vs<br>HC | PCA vs<br>tAD | PCA vs<br>lvPPA | tAD vs<br>lvPPA | HC vs<br>tAD | HC vs<br>lvPPA |
| --- | --- | --- | --- | --- | --- | --- | --- | --- | --- | --- |
| **Fluency** |  |  |  |  |  |  |  |  |  |  |
| **Category Fluency** | 37.0(8.6)<br>)<br>n=165 | 19.1(9.6)<br>)<br>n=99 | 19.1(8.4)<br>)<br>n=105 | 15.3(8.4)<br>)<br>n=107 | <.0001 | 0.981 | 0.007 | 0.029 | <.0001 | <.0001 |
| **Verbal Fluency: Phonemic Test** | 29.4(8.4)<br>)<br>n=165 | 20.3(10.8)<br>)<br>n=93 | 17.8(9.4)<br>)<br>n=105 | 15.3(8.0)<br>)<br>n=101 | <.0001 | 0.481 | 0.002 | 0.095 | <.0002 | <.0003 |
| **Reading** |  |  |  |  |  |  |  |  |  |  |
| **Regular word reading** | 15.0(0.2)<br>)<br>n=165 | 14.1(2.0)<br>)<br>n=20 | 14.9(0.7)<br>)<br>n=105 | 14.3(1.5)<br>)<br>n=32 | <.0001 | 0.001 | 0.689 | 0.007 | 0.686 | 0.000 |
| **Irregular word reading** | 14.6(0.8)<br>)<br>n=165 | 12.8(2.8)<br>)<br>n=20 | 13.7(1.8)<br>)<br>n=105 | 11.1(2.9)<br>)<br>n=32 | <.0001 | 0.054 | 0.004 | <.0001 | 0.000 | <.0001 |
| **Sentence reading** | 4.8(0.5)<br>)<br>n=165 | 3.9(1.9)<br>)<br>n=18 | 4.6(0.7)<br>)<br>n=105 | 3.2(1.5)<br>)<br>n=27 | <.0001 | 0.003 | 0.024 | <.0001 | 0.179 | <.0001 |
| **Semantics** |  |  |  |  |  |  |  |  |  |  |
| **Semantic Word-picture matching test** | 19.9(1.6)<br>)<br>n=165 | 18.0(3.0)<br>)<br>n=21 | 19.6(2.0)<br>)<br>n=105 | 19.7(0.8)<br>)<br>n=32 | 0.000 | 0.002 | 0.008 | 0.998 | 0.761 | 0.968 |
| **Semantic Associates Test** | 16.0(0.2)<br>)<br>n=165 | 15.0(2.2)<br>)<br>n=18 | 15.2(1.7)<br>)<br>n=105 | 15.4(1.2)<br>)<br>n=29 | 0.006 | 0.006 | 0.591 | 0.742 | <.0001 | 0.150 |
| **Semantic Associates Test - Total correct animal associations** | 8.0(0.2)<br>)<br>n=165 | 7.3(1.6)<br>)<br>n=18 | 7.6(0.9)<br>)<br>n=105 | 7.7(0.7)<br>)<br>n=29 | 0.002 | 0.506 | 0.281 | 0.820 | 0.000 | 0.338 |
| **Semantic Associates Test - Total correct tool associations** | 8.0(0.1)<br>)<br>n=165 | 7.7(0.7)<br>)<br>n=18 | 7.6(0.9)<br>)<br>n=105 | 7.7(0.8)<br>)<br>n=29 | 0.132 | 0.982 | 0.987 | 0.783 | <.0001 | 0.145 |
| **Repetition** |  |  |  |  |  |  |  |  |  |  |
| **Sentence repetition** | 4.4(0.8)<br>)<br>n=165 | 3.9(1.4)<br>)<br>n=21 | 4.0(0.9)<br>)<br>n=105 | 2.3(1.6)<br>)<br>n=25 | 0.164 | 0.965 | <.0001 | <.0001 | 0.023 | <.0001 |
| **Number Span Test: Forward — Number of correct trials** | 8.5(2.6)<br>)<br>n=165 | 6.3(2.3)<br>)<br>n=102 | 6.5(2.1)<br>)<br>n=104 | 3.7(2.0)<br>)<br>n=111 | <.0001 | 0.784 | <.0001 | <.0001 | <.0001 | <.0001 |
| **Naming** |  |  |  |  |  |  |  |  |  |  |
| **Noun naming** | 15.9(0.4)<br>)<br>n=165 | 13.4(2.3)<br>)<br>n=18 | 15.5(1.2)<br>)<br>n=105 | 13.6(3.1)<br>)<br>n=27 | <.0001 | <.0001 | 0.913 | <.0001 | 0.097 | <.0001 |
| **Verb naming** | 15.9(0.5<br>)<br>n=165 | 10.1(4.7<br>)<br>n=18 | 14.9(1.6<br>)<br>n=105 | 12.4(3.3<br>)<br>n=25 | <.0001 | <.0001 | <.0001 | <.0001 | 0.001 | <.0001 |
| **Multilingual**<br>**Naming task** | 30.3(1.8<br>)<br>n=165 | 23.7(6.8<br>)<br>n=79 | 27.3(4.4<br>)<br>n=105 | 19.6(8.7<br>)<br>n=100 | <.0001 | 0.000 | <.0001 | <.0001 | 0.000 | <.0001 |

**Table 3.**
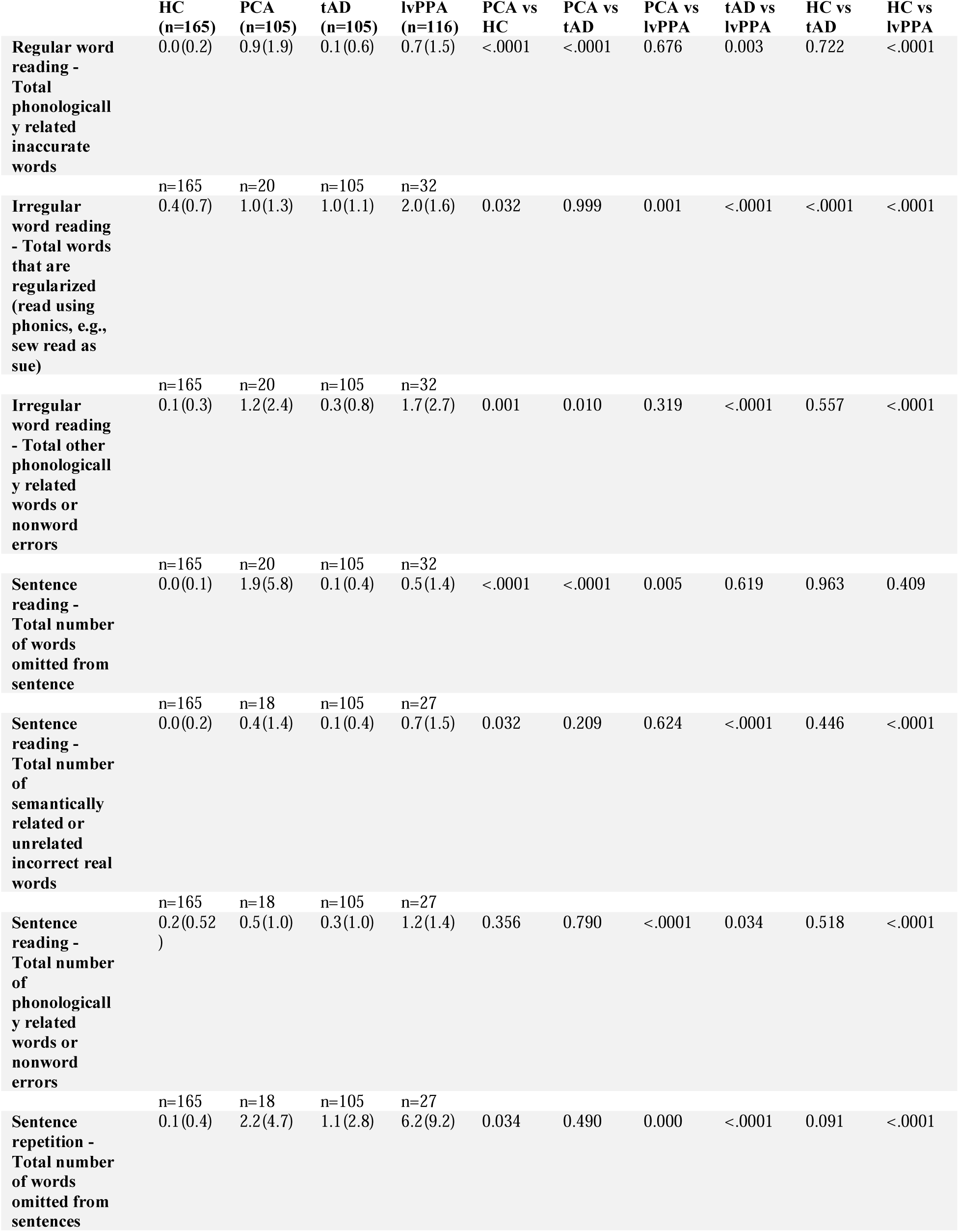
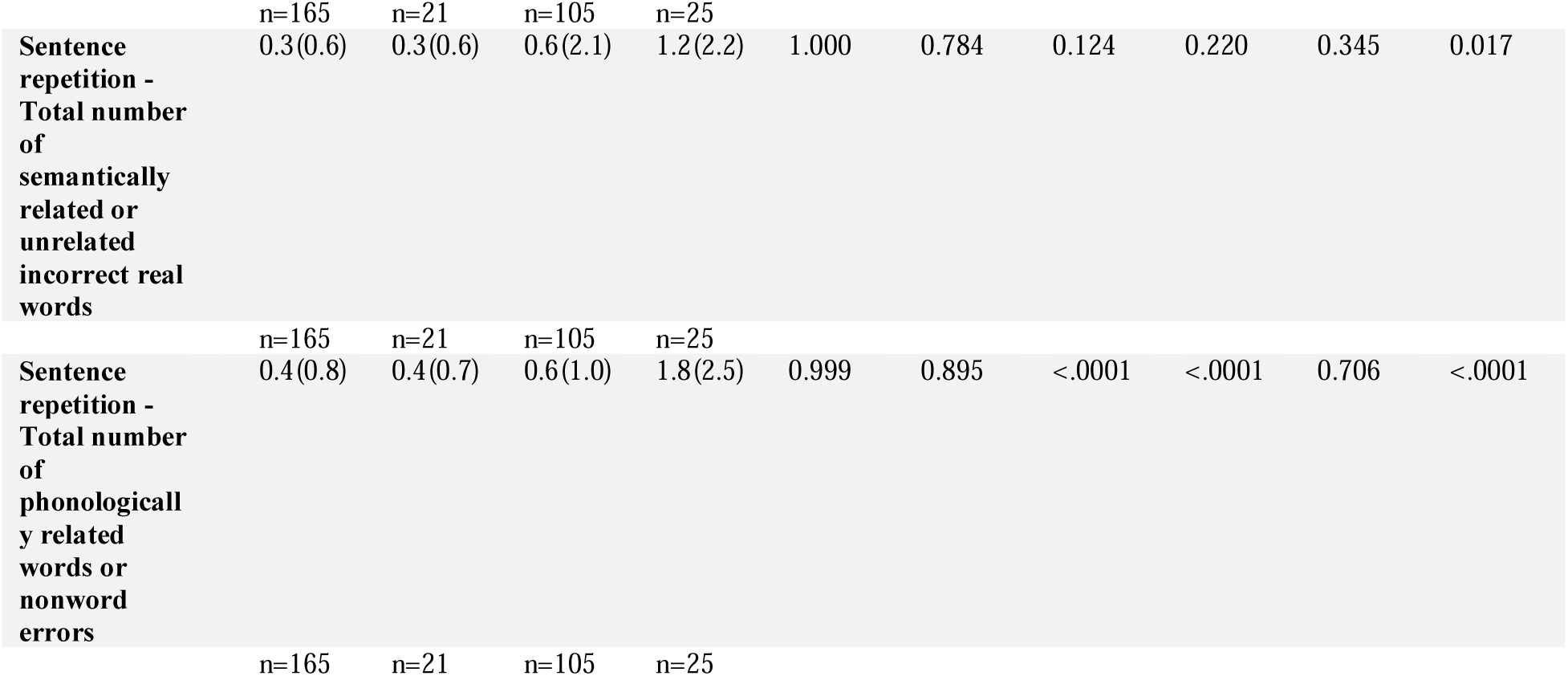
Between-group comparisons on language tests error scores.

#### 3.2.1. Fluency

PCA patients performed worse than HC in both category and phonemic fluency tasks (p < .0001; Figure 1). When comparing PCA with tAD, no significant differences were observed in performance on either task (p = .981 on category; p = .481 on phonemic). In contrast, patients with lvPPA demonstrated significantly poorer performance than PCA in both category (p < .01) and phonemic fluency (p < .01).

**Figure 1.**
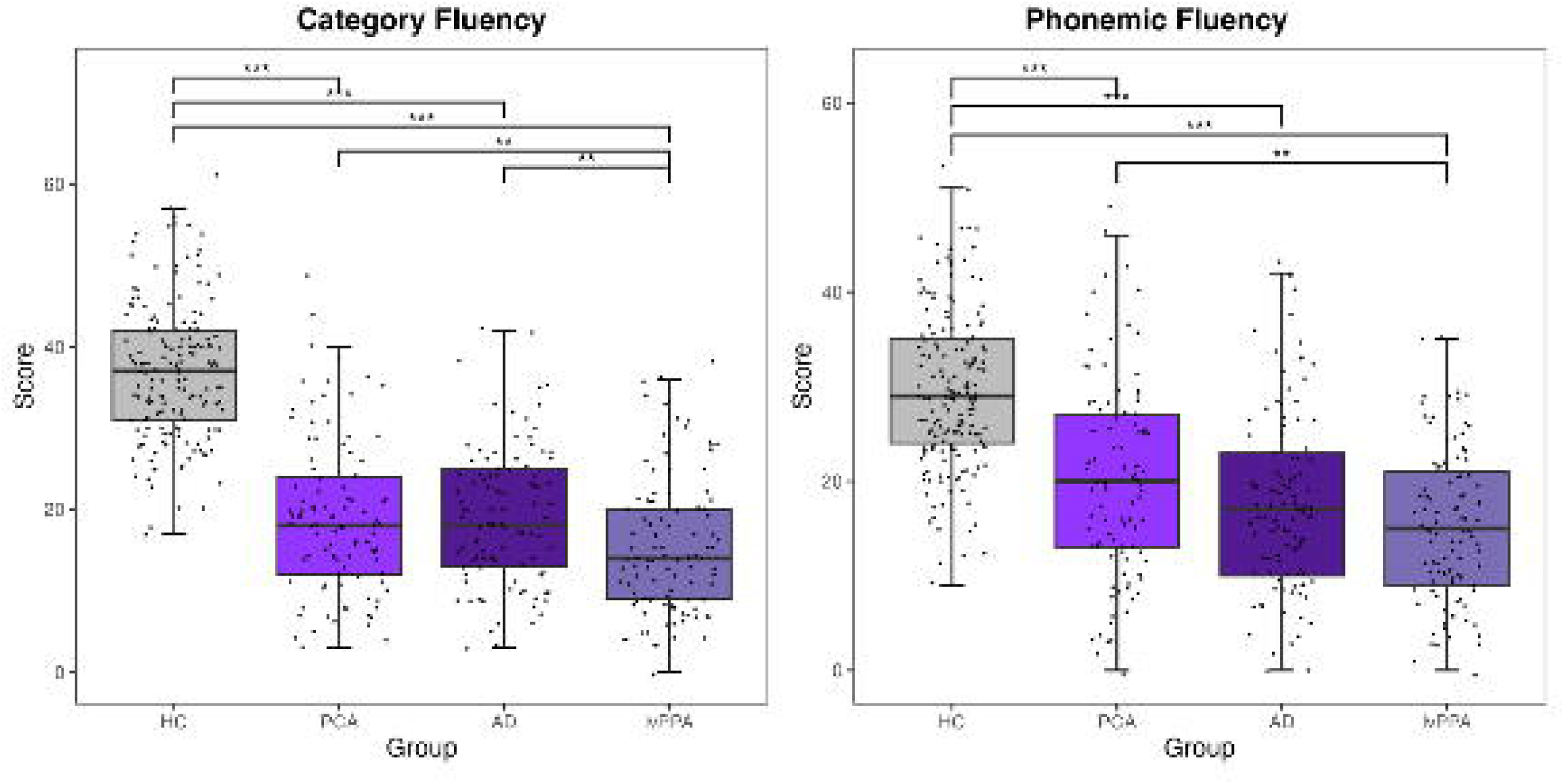
Comparison of verbal fluency performance between PCA, lvPPA, tAD patients and HC. ANCOVAs controlling for age, sex and education were conducted (* p < 0.05; ** p < 0.01; *** p < 0.001)

#### 3.2.2. Reading

In all three reading assessments, individuals with PCA had significantly lower scores than HC (p < .0001; Figure 2). In the regular word reading task, PCA’s performance was significantly different from tAD (p < .0001) but not from lvPPA (p = .689). In the irregular word reading task, PCA participants outperformed those with lvPPA (p < .01), but there was no difference between PCA and tAD (p=.054). In the sentence reading task, PCA patients scored worse than tAD patients (p < .01) but scored higher than lvPPA patients (p < .05).

**Figure 2.**
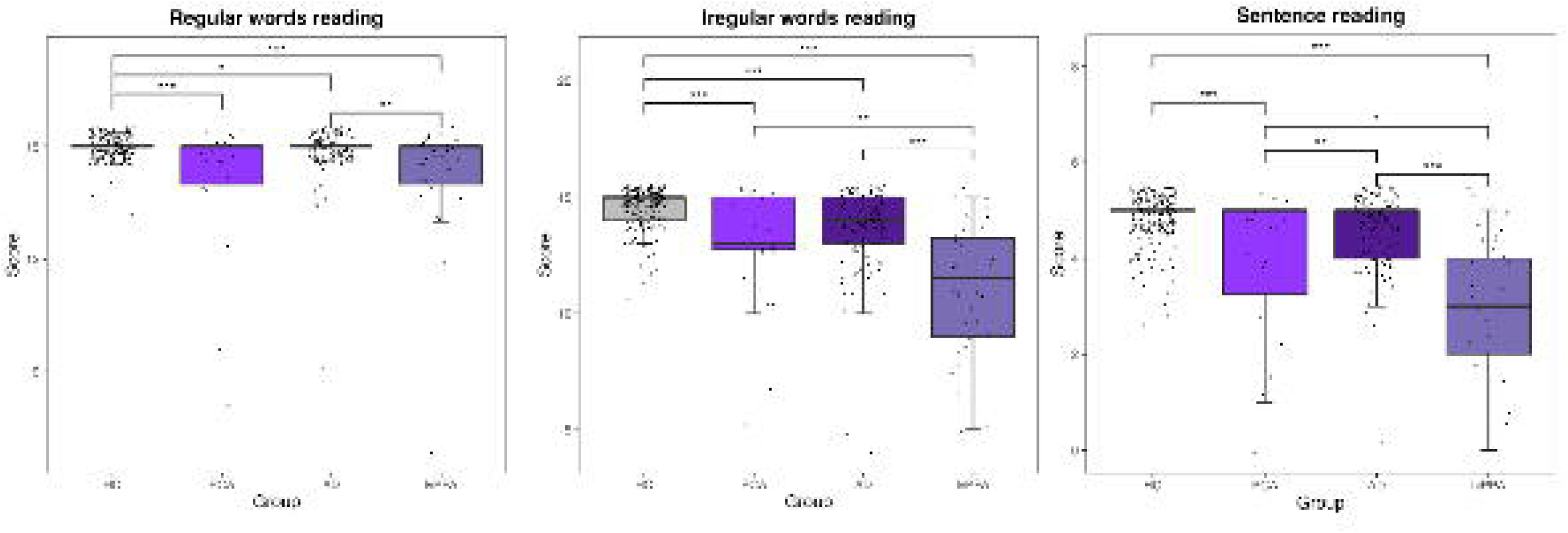
Comparison of reading performance between PCA, lvPPA, tAD patients and HC. ANCOVAs controlling for age, sex and education were conducted (* p < 0.05; ** p < 0.01; *** p < 0.001)

When looking at the interaction between group (HC vs PCA) and stimuli type (regular vs irregular words), the results indicated a significant interaction (F (1, 366) = 8.9, p < .001; Figure 6A). We conducted independent t-tests comparing irregular and regular word reading for the PCA and HC groups, respectively. Both groups showed better performance on regular word reading compared to irregular word reading (p = .001 for PCA; p < .0001 for HC), although the interaction effect suggest that the stimuli type effect was more significant in PCA patients.

Regarding error profiles in regular word reading, PCA patients made significantly more phonological errors than both the tAD and HC groups (p < .0001 for both), but not compared to lvPPA (p = .676). For irregular word reading, regularization errors in the PCA group were similar to tAD (p = .999), but significantly fewer than lvPPA (p < .001), and more frequent than HC (p < .05). Additionally, PCA patients made more phonological errors than the HC (p < .001) and tAD group (p < .05) and, but not compared to lvPPA (p = .319). In sentence reading, omission errors were most prominent in PCA, with significantly more frequent errors compared to HC and tAD (p < .0001) and lvPPA (p < .01). Semantic errors were significantly different between PCA and HC (p < .05), but no significant differences were observed between PCA and tAD (p = .209) or lvPPA (p = .624). Phonological errors did not differ significantly between PCA and HC or tAD, but PCA patients made significantly fewer phonological errors than lvPPA (p < .0001; Figure 7).

#### 3.2.3. Semantics

In the semantic word-picture matching test, PCA patients had significantly lower scores compared to HC (p < .0001), tAD (p < .01), and lvPPA (p < .01; Figure 3). In the semantic association task, PCA patients also performed worse than HC (p < .01) and tAD (p < .01), but no significant differences were found when comparing PCA to lvPPA (p = .591; Figure 3).

**Figure 3.**
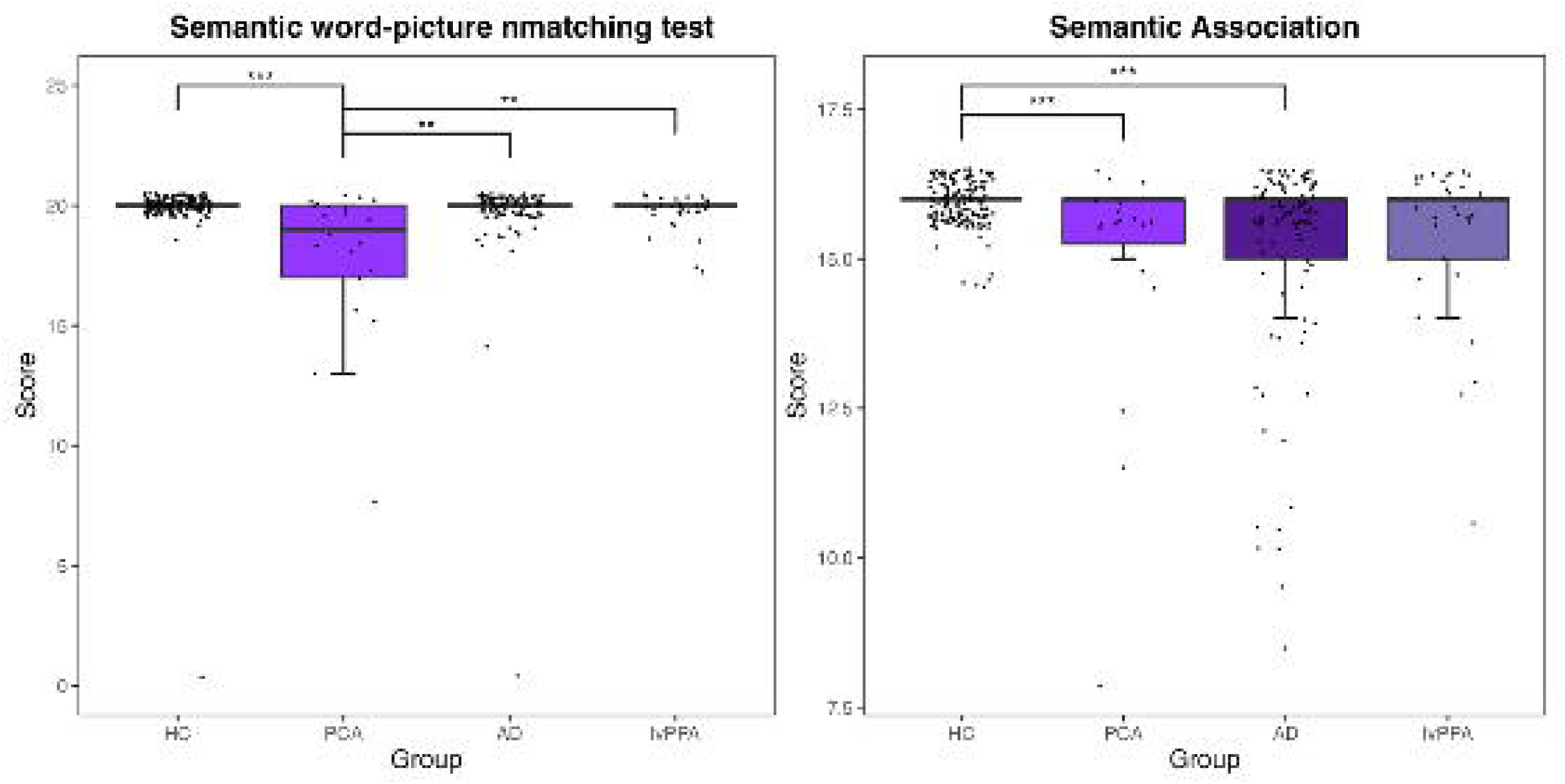
Comparison of semantic performance between PCA, lvPPA, tAD patients and HC. ANCOVAs controlling for age, sex and education were conducted (* p < 0.05; ** p < 0.01; *** p < 0.001)

#### 3.2.4. Short term verbal memory

In the sentence repetition task, there was a trend for lower performance in PCA patients compared to HC, but this difference did not reach statistical significance (p = .164; Figure 4). However, in the digit forward span task, PCA patients performed significantly worse than HC (p < .0001). When comparing PCA with tAD, no significant differences were found in either sentence repetition (p = .965) or digit forward span (p = .784). In contrast, lvPPA patients showed significantly worse performance compared to PCA in both sentence repetition (p < .0001) and digit forward span (p < .0001).

**Figure 4.**
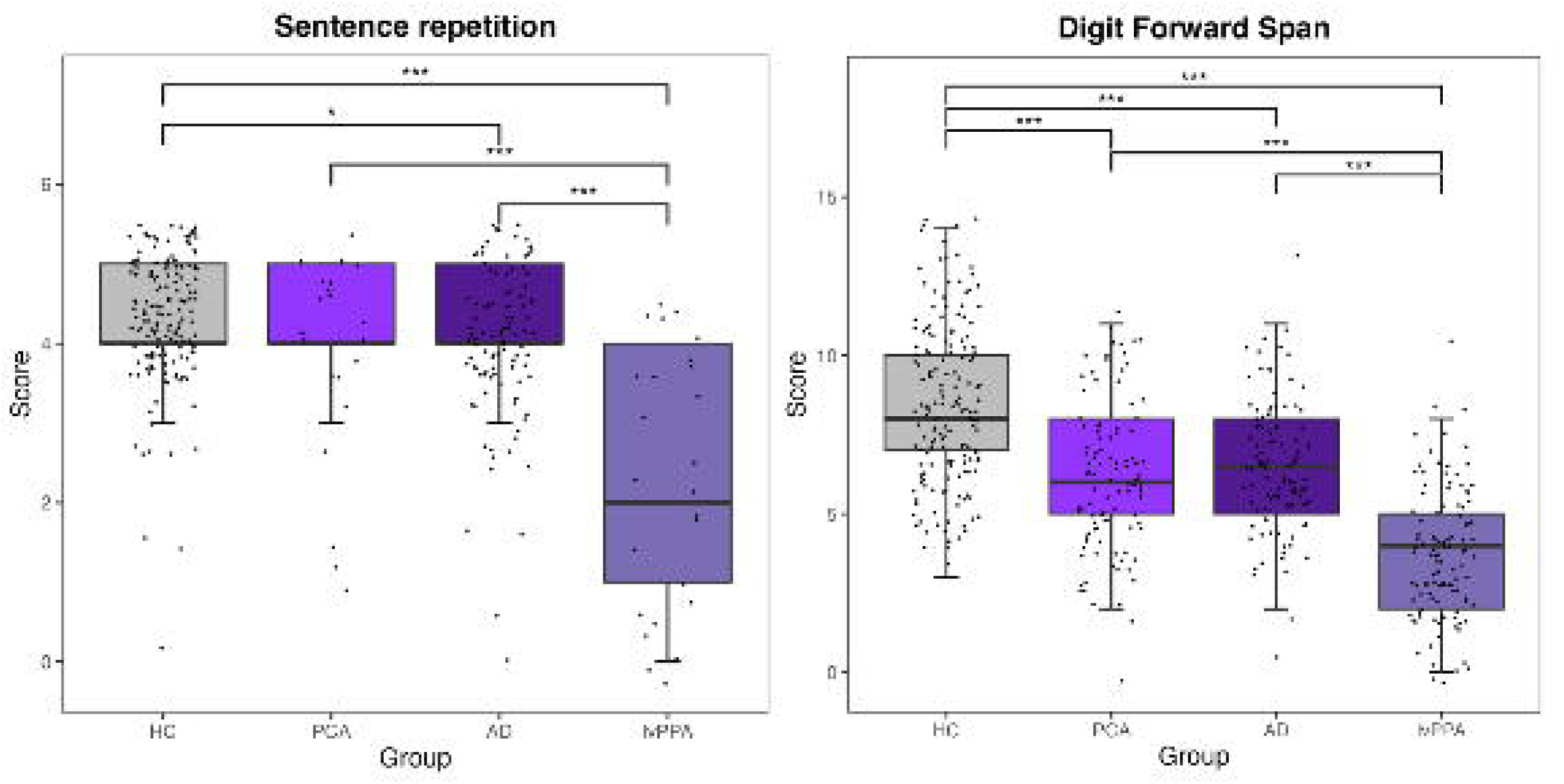
Comparison of verbal short term memory performance between PCA, lvPPA, tAD patients and HC. ANCOVAs controlling for age, sex and education were conducted (* p < 0.05; ** p < 0.01; *** p < 0.001)

Regarding errors in sentence repetition, PCA patients made more omission errors compared to HC (p < .05) and significantly fewer omission errors than lvPPA patients (p < .0001), but the difference with tAD was not statistically significant (p = .527). In terms of semantic errors, PCA patients performed similarly to other groups (p = 1.000 with HC; p = .784 with tAD; p = .124 with lvPPA. For phonological errors, PCA patients showed similar performance to HC (p = .999) and tAD (p = .895) but significantly fewer numbers than lvPPA (p < .0001).

#### 3.2.5. Naming

Compared to HC, PCA patients exhibited worse performance across all three naming tasks (p < .0001; Figure 5). In the Multilingual Naming Test, PCA patients performed worse than tAD (p = .0001), but better than lvPPA (p < .0001). In noun and verb naming, PCA performed worse than both HC and tAD (p < .0001). PCA performed worse than lvPPA in verb naming (p < .0001), but similarly in noun naming (p = .913).

**Figure 5.**
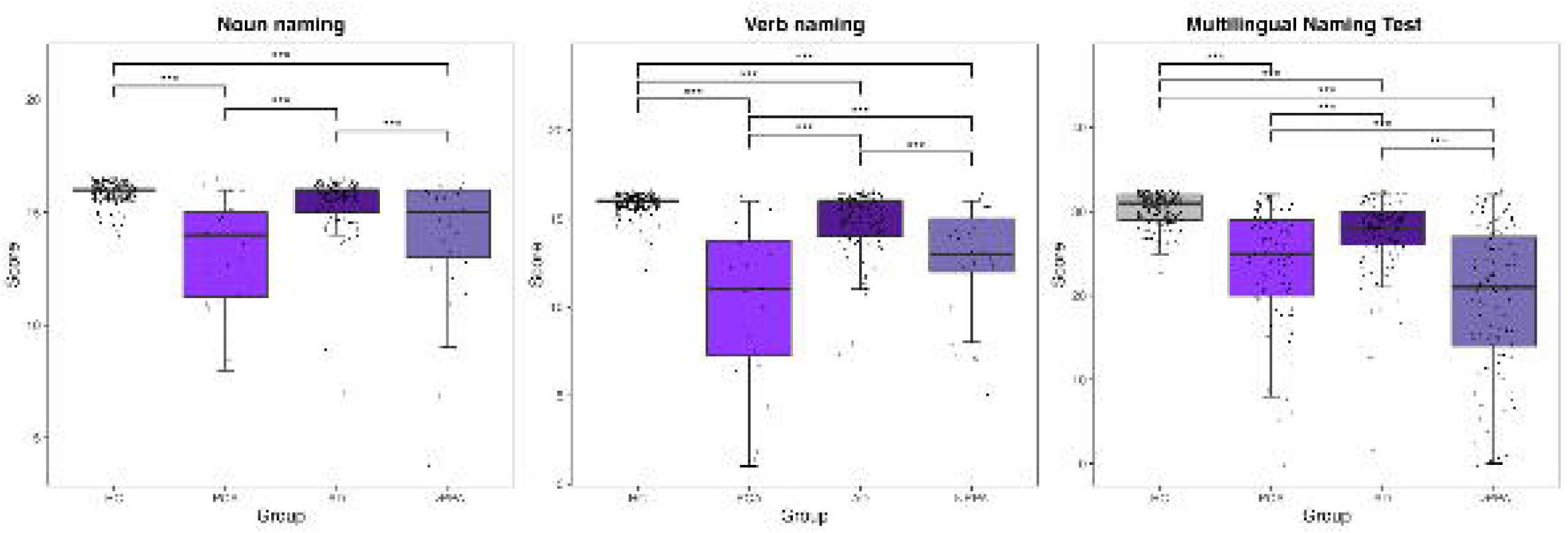
Comparison of confrontation naming performance between PCA, lvPPA, tAD patients and HC. ANCOVAs controlling for age, sex and education were conducted (* p < 0.05; ** p < 0.01; *** p < 0.001)

When analyzing the interaction between group (HC vs PCA) and stimuli type (noun vs verb naming), a significant interaction effect was found (F(1, 362) = 60.9, p < .0001; Figure 6C). Independent t-tests comparing noun and verb naming for the PCA and HC groups showed that PCA performed better on noun naming compared to verb naming (p = .001), while HC performed similarly on the two tasks (p = .477).

**Figure 6.**
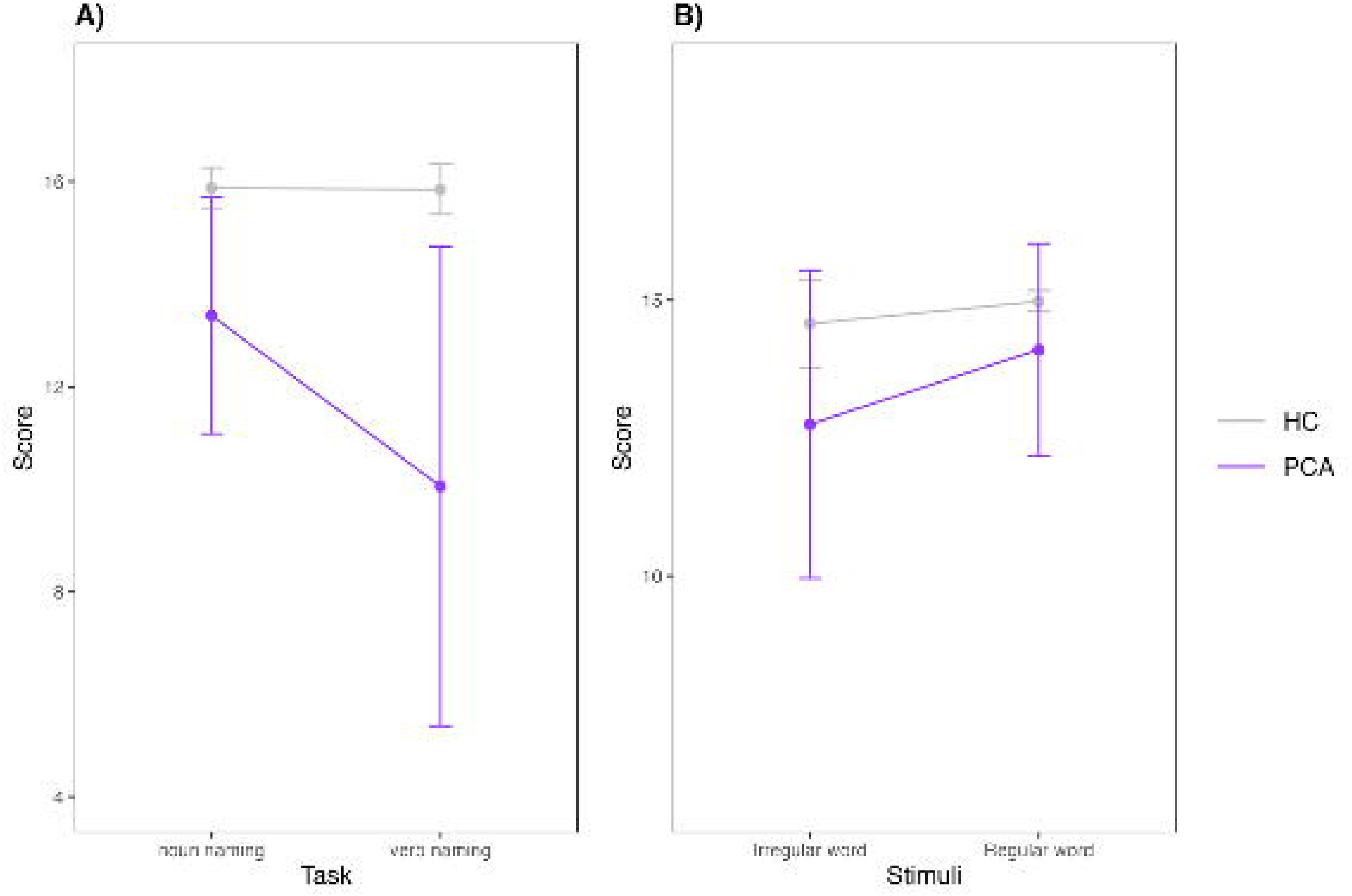
Group-condition interactions for three tasks: A) Performance on naming nouns vs verbs, in HC and PCA patients, in the Noun and verb naming task; B) Performance on irregular vs regular word reading, in HC and PCA patients, from the noun reading test; Two-way ANCOVAs were conducted controlling for age, sex, education).

**Figure 7.**
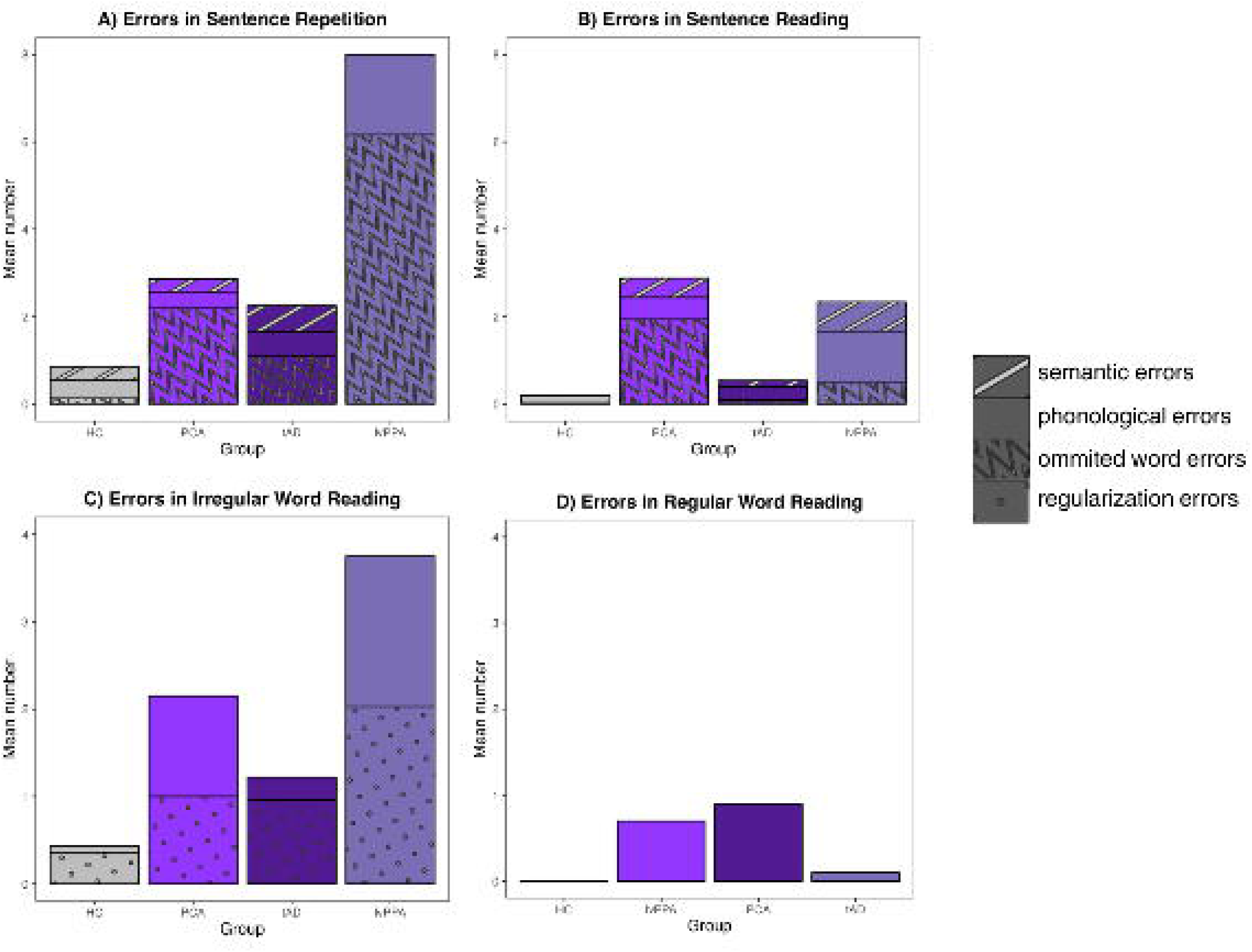
Mean number of errors by type and by group for 4 tasks: A) Sentence repetition; B) Sentence reading; C) Irregular word reading; D) Regular word reading.

## 4. Discussion

The study aimed to unravel the language profile in PCA patients. Our findings revealed a global decline in visual and non-visual language functions among PCA patients compared to HC, with no spared domains. Furthermore, we investigated specific language errors in reading and sentence repetition, and we found that PCA patients committed a mix of phonological, semantic and word omission errors. Additionally, we compared PCA with tAD and lvPPA, and our results suggest that overall, the language impairments observed follow a tAD > PCA > lvPPA pattern. While PCA is primarily characterized by deficits in visual and visuospatial functions, our study emphasizes the significance of investigating impairments in cognitive domains other than the visual domain.

Unsurprisingly, we found significant impairments in visual language tasks, in PCA, including reading, semantics, and naming. These results are in line with the diagnostic criteria^1,2,3^ and previous research on alexia, agraphia and visual agnosia in PCA^14,15,2^. Nonetheless, the current study attempted to go beyond visual language impairments using three different approaches. First, we investigated language performance in tasks that are independent of visual functions. This allowed us to show that even in the early stages of the PCA, impairments in short term verbal memory and verbal fluency are evident, suggesting broader phonological loop, lexicon size, semantic retrieval, and executive functioning impairments. Interestingly, the digit span forward task appeared more sensitive than the FTLD-Module sentence repetition task in identifying short-term verbal memory deficits in PCA. It is possible that the acalculia observed in PCA patients contributes to impaired performance in repeating numbers specifically^2^. Moreover, the digit span task requires short-term memorization of random numbers, with no opportunity to draw on semantic knowledge or context. In contrast, the sentence repetition task sometimes allows patients to rely on semantic cues. This reliance on semantic memory could potentially mask the severity of their short-term verbal memory deficits when performing the sentence repetition task.

Our second approach was to look into the errors profile in language tasks in PCA, which can help in clarifying whether the impaired performance is due to visual or non-visual errors. In that regard, we found a mixed pattern of errors in PCA. They showed a high frequency of word omission errors when reading sentences, which indicate early visual processing deficits. However, both phonological and semantic (regularization) errors were also observed in word reading, which suggests that reading impairments might be multifaceted in PCA.

Finally, our last approach was to investigate within-task dissociations based on the stimuli type, while controlling for the task at hand which remains the same. First, a noun-verb dissociation was observed in PCA, with better performance in nouns naming vs verb naming, while this dissociation was not found in HC. In a previous study, it has been shown that impaired noun naming was associated with atrophy in the left anterior temporal regions, while impaired verb naming was associated with atrophy in the left inferior parietal lobe, posterior middle temporal gyrus, and inferior and middle frontal gyrus^43^. The atrophy pattern in PCA shows more overlap with regions related to verbs than nouns, which may explain such dissociation. It’s noticeable that the noun-verb dissociation was not found in patients with lvPPA, making this dissociation in PCA valuable for differential diagnosis. In lvPPA, noun and verb deficits are more evenly distributed^43,44^, further highlighting the specificity of this dissociation in PCA. Secondly, PCA patients exhibited a dissociation between irregular word reading to regular word reading, with worse performance in irregular words reading. Since regular words primarily rely on phonological decoding and irregular words involve semantic processing, this indicates semantic processing deficits also occur on PCA.

While our study partially supports the notion that PCA patients can exhibit a logopenic-like language profile (decline in naming, sentence repetition and phonological errors), it also highlighted more complex nuances that can contribute to the differential diagnosis of PCA and lvPPA. First, we found that language impairments in PCA are generally less severe: PCA patients performed better than lvPPA patients in verbal fluency (both category and phonemic). Such difference may be mediated by more pronounced deficits in executive functioning in lvPPA, which are critical for retrieving words during fluency tasks. PCA is also better on verb naming, and sentence repetition, and they made fewer phonological errors in irregular word reading. Additionally, the severity of the sentence repetition deficits was actually comparable to those found in tAD, which suggest that they may not be as distinctive or defining as previously thought in PCA. Our findings also indicate that the language impairments in PCA extended beyond the lvPPA core criteria. For instance, PCA patients performed worse on the single-word comprehension task (which may depends on visual impairment), whereas lvPPA patients showed relatively intact performance in these areas. This suggests that semantic and visuospatial deficits in PCA contribute to language impairments not typically observed in lvPPA. However, given assessment of semantic abilities in PCA was conducted through visually based tasks, it is possible that the observed impairments are attributable to visual processing difficulties. Nonetheless, the occurrence of regularization errors in PCA patients offers some indication that semantic deficits may indeed be present. One possible explanation for the semantic deficits in PCA patients is that damage to the occipito-parietal regions impairs their ability to form and manipulate mental images. This disruption in visual imagery can make it harder for them to access and retrieve the meanings of objects, leading to semantic difficulties. The link between mental imagery and semantic deficits arises because both processes rely on shared neural networks. When visual imagery fails, access to mental representations of objects is disrupted, weakening semantic knowledge.^45,46^. However, this hypothesis requires further investigation through the use of non-visual semantic tasks, to determine whether the semantic deficits in PCA patients persist in the absence of visual stimuli.

Previous research has often suggested that the language profiles of PCA and tAD are largely comparable^2,47^. However, our findings indicate that language impairments in PCA are more pronounced when visual components are involved, especially in reading, single-word comprehension and naming. Furthermore, PCA patients made more omission errors than tAD patients during sentence reading and more phonological errors in irregular word reading than tAD patients, indicating that PCA patients struggle more with processing phonological components of visually presented words. Our results suggest that these tests could be helpful in the differential diagnosis between PCA and tAD.

This study has a few limitations. Despite having a large sample size, we encountered missing data, specifically regarding the FTLD-MOD language tasks. In addition, visual language tasks are suboptimal for assessing linguistic abilities for PCA patients. Future research should focus on non-visual language tasks, such as oral spelling, auditory naming, and auditory semantic judgment of word definitions, to better assess language impairments in PCA. Additionally, grammar, motor speech, and prosody should be investigated, as these areas were not covered in our study but are essential for a comprehensive understanding of PCA’s language profile. It is also crucial to explore the real-life impact of language difficulties in PCA patients through discourse analysis and real-life communication questionnaires. These approaches can provide insights into how language impairments affect daily functioning and quality of life, complementing the information gained from traditional neuropsychological assessments. Finally, not all PCA patients included in the study had confirmed AD biomarkers.

In conclusion, the manifestation of both visual and non-visual language impairments in PCA, even in its early stages, underscores the complexity of this neurodegenerative condition. Our findings highlight that phonological errors and word omission errors play predominant roles in the failure of language tasks in PCA, suggesting underlying deficits in phonological processing, while semantic deficits are less significant. PCA demonstrates relatively preserved linguistic function compared to lvPPA, distinguishing between PCA and typical tAD based solely on linguistic assessment is difficult. These insights emphasize the need for comprehensive evaluations that consider both visual and non-visual language abilities to accurately diagnose and manage individuals with PCA.

## Data Availability Statement

The data used in this study are available from the National Alzheimer’s Coordinating Center (NACC) database: https://naccdata.org/requesting-data/submit-data-request

## Funding Information Authors’ contributions

Linshan Wang and Maxime Montembeault designed the study. Linshan Wang analyzed the data, interpreted the results, and wrote the manuscript. All authors read, edited, and approved the final manuscript.

## Acknowledgements

The NACC database is funded by NIA/NIH Grant U24 AG072122. NACC data are contributed by the NIA-funded ADRCs: P30 AG062429 (PI James Brewer, MD, PhD), P30 AG066468 (PI Oscar Lopez, MD), P30 AG062421 (PI Bradley Hyman, MD, PhD), P30 AG066509 (PI Thomas Grabowski, MD), P30 AG066514 (PI Mary Sano, PhD), P30 AG066530 (PI Helena Chui, MD), P30 AG066507 (PI Marilyn Albert, PhD), P30 AG066444 (PI John Morris, MD), P30 AG066518 (PI Jeffrey Kaye, MD), P30 AG066512 (PI Thomas Wisniewski, MD), P30 AG066462 (PI Scott Small, MD), P30 AG072979 (PI David Wolk, MD), P30 AG072972 (PI Charles DeCarli, MD), P30 AG072976 (PI Andrew Saykin, PsyD), P30 AG072975 (PI David Bennett, MD), P30 AG072978 (PI Neil Kowall, MD), P30 AG072977 (PI Robert Vassar, PhD), P30 AG066519 (PI Frank LaFerla, PhD), P30 AG062677 (PI Ronald Petersen, MD, PhD), P30 AG079280 (PI Eric Reiman, MD), P30 AG062422 (PI Gil Rabinovici, MD), P30 AG066511 (PI Allan Levey, MD, PhD), P30 AG072946 (PI Linda Van Eldik, PhD), P30 AG062715 (PI Sanjay Asthana, MD, FRCP), P30 AG072973 (PI Russell Swerdlow, MD), P30 AG066506 (PI Todd Golde, MD, PhD), P30 AG066508 (PI Stephen Strittmatter, MD, PhD), P30 AG066515 (PI Victor Henderson, MD, MS), P30 AG072947 (PI Suzanne Craft, PhD), P30 AG072931 (PI Henry Paulson, MD, PhD), P30 AG066546 (PI Sudha Seshadri, MD), P20 AG068024 (PI Erik Roberson, MD, PhD), P20 AG068053 (PI Justin Miller, PhD), P20 AG068077 (PI Gary Rosenberg, MD), P20 AG068082 (PI Angela Jefferson, PhD), P30 AG072958 (PI Heather Whitson, MD), P30 AG072959 (PI James Leverenz, MD)

